# Automated Versus Conventional Annular Suturing in Totally Endoscopic Aortic Valve and Ascending Aorta Replacement: A Prospective Randomized Trial

**DOI:** 10.64898/2026.10.27.26366669

**Authors:** Marwan Hamiko, Julia Rogaczewski, Saad Salamate, Ali Bayram, Florian Piekarski, Kaveh Eghbalzadeh, MIriam Silaschi, Jacquline Kruse, Lazar Velicki, Michael E. Ibrahim, Ali El-Sayed Ahmad, Farhad Bakhtiary

## Abstract

**Background:** Totally endoscopic aortic valve replacement (AVR) combined with ascending aorta replacement (AAR) remains technically demanding, with prolonged CPB and ischemic times. Annular suturing is a principal technical limitation; automated devices aim to simplify this step and improve efficiency. This study compared early clinical outcomes between automated and conventional annular suturing in totally endoscopic AVR with concomitant AAR.

**Methods:** Between August 2023 and February 2026, 86 patients undergoing totally endoscopic AVR with concomitant AAR via right anterior minithoracotomy (RAMT) were randomized 1:1 to automated annular (RAM group; n=43) or conventional suturing (conventional group; n=43) in this single-center trial. Primary endpoints were technical success and procedural times; secondary endpoints included complications and 30-day mortality.

**Results:** Baseline characteristics were comparable, bicuspid aortic valve morphology was more frequent in the RAM group (74.4% vs. 53.5%; P = 0.043). Automated suturing was associated with shorter median procedure (120.8 vs. 144.0 minutes; P = 0.004), CPB (90.0 vs. 105.0 minutes; P = 0.005), and aortic cross-clamp times (62.0 vs. 74.0 minutes; P = 0.003), respectively. No stroke, conversion to sternotomy, paravalvular leakage, or 30-day mortality occurred in either group. Hospital stay was shorter with automated suturing (6.0 vs. 7.0 days; P = 0.037), while total treatment cost, accounting for incremental device cost, did not differ significantly (P = 0.421).

**Conclusions:** Automated annular suturing appears to be a safe, efficient alternative to conventional suturing in totally endoscopic AVR with AAR, associated with reduced operative and ischemic times and shorter hospital stay without compromising early outcomes.

**CLINICAL PERSPECTIVE:** *What Is New?:* In this randomized trial of totally endoscopic aortic valve replacement with ascending aorta replacement (n=86), automated annular suturing with the RAM device was associated with shorter procedure, cardiopulmonary bypass, and aortic cross-clamp times and a shorter hospital stay (6 vs 7 days) than conventional suturing. Earlier evidence for automated annular suturing came mainly from observational series in isolated valve replacement, and this trial extends the observation to combined aortic valve and ascending aorta surgery. No stroke, conversion to sternotomy, paravalvular leakage, or 30-day mortality occurred in either group, and total treatment cost did not differ significantly because the incremental device cost offset the savings from the shorter hospital stay.

*What Are the Clinical Implications?:* Automated annular suturing may be a safe and efficient option to reduce ischemic and bypass times in technically demanding totally endoscopic aortic procedures at experienced centers. Centers considering adoption should expect cost neutrality rather than cost savings during the index hospitalization. Larger multicenter trials with longer follow-up, including centers earlier in their learning curve, are needed to confirm generalizability and valve durability.

**GRAPHICAL ABSTRACT:** 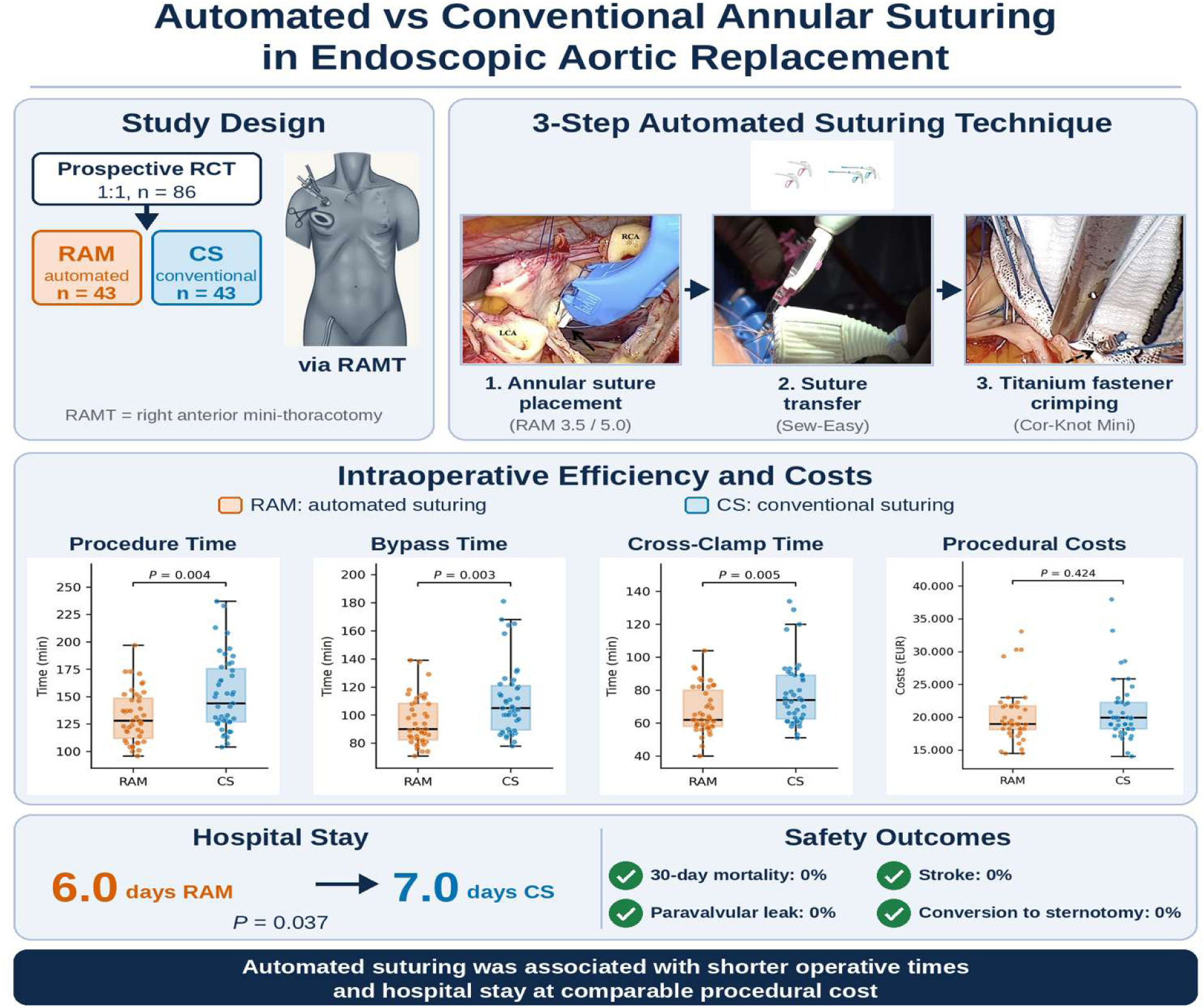

## INTRODUCTION

Minimally invasive and totally endoscopic approaches in cardiac surgery have evolved substantially over the past decade, enabling increasingly complex procedures to be performed through limited access incisions while maintaining excellent clinical outcomes. Endoscopic aortic valve replacement (AVR) through right anterior minithoracotomy (RAMT) has emerged as an effective alternative to conventional sternotomy, offering reduced surgical trauma, faster postoperative recovery, shorter hospitalization, and improved patient satisfaction.^1–3^

Despite these advantages, totally endoscopic procedures remain technically demanding. One of the principal limitations is the placement of annular sutures within a restricted operative field under endoscopic visualization. This challenge becomes even more pronounced in combined procedures involving replacement of the ascending aorta (AAR), where prolonged cardiopulmonary bypass (CPB) and aortic cross-clamp times may negatively affect postoperative outcomes.^4–5^

To address these limitations, automated annular suturing devices have recently been introduced into minimally invasive valve surgery.^6–7^ The RAM automated suturing device (LSI solutions, Victor, NY, USA) was developed to standardize annular suture placement and facilitate reproducible implantation of prosthetic valves during totally endoscopic procedures. Previous observational studies demonstrated promising results regarding procedural efficiency and safety in isolated endoscopic AVR.^1,8^

However, evidence regarding the use of automated suturing in more complex procedures such as combined AVR and AAR remains limited. The current prospective randomized trial therefore aimed to compare the clinical feasibility and efficacy of automated annular suturing using the RAM device with conventional manually placed annular sutures in patients undergoing totally endoscopic AVR with concomitant AAR.

## MATERIAL AND METHODS

### Study Design

Following approval by the institutional ethics committee of the University of Bonn (approval no. 184/23; August 7, 2023) and acquisition of written informed consent from all participants, patients were prospectively enrolled between August 2023 and February 2026 in this single-center, randomized trial (German Registry for Clinical Studies, Trial number: DRKS00030520, registry date: 18^th^ December 2023). All patients were scheduled to undergo totally endoscopic minimally invasive AVR with concomitant AAR via RAMT and were allocated in a 1:1 ratio to either automated annular suturing with the RAM device or conventional, manually placed pledgeted annular sutures. Treatment allocation was determined by a computer-generated randomization sequence established prior to the first enrollment.

### Patient Selection and Study Groups

Patients with aortic valve pathology (stenosis or regurgitation) and a concomitant aneurysm of the ascending aorta (AA) or aortic root (AR) meeting current ESC/EACTS criteria for surgical intervention (diameter ≥45 mm) were screened for eligibility for a totally endoscopic RAMT approach.^9^

Mandatory preoperative anatomical assessment consisted of contrast-enhanced computed tomography (CT) scan, analyzed with 3mensio software (Pie Medical Imaging, Bilthoven, The Netherlands), systematically evaluating aortic annulus and sinotubular junction diameters, maximum AR and AA diameters, coronary ostium height, spatial relationship between the aortic axis and anterior chest wall, optimal intercostal access, and femoral/axillary vessel suitability for CPB cannulation.

Inclusion criteria comprised a distal ascending aortic diameter ≤ 42 mm, maintained along an adequate distal anastomotic cuff of at least 2 cm to allow safe cross-clamping: absence of significant calcification or kinking of the thoracic aorta and peripheral access vessels. This threshold was established through our institutional experience, where diameters exceeding 42 mm were consistently associated with attenuated aortic wall tissue and increased anastomotic bleeding risk.

Exclusion criteria were partial or complete sternotomy, concomitant cardiac procedures, acute or chronic aortic dissection, planned arch replacement (hemiarch or total arch), active infective endocarditis, and redo cardiac surgery.

Baseline characteristics included cardiovascular risk factors, relevant comorbidities, echocardiographic and CT findings, and New York Heart Association (NYHA) functional class. Perioperative and postoperative data were obtained from institutional electronic medical records, and preoperative risk was assessed using European System for Cardiac Operative Risk Evaluation (EuroSCORE II).

Of patients screened for feasibility of a totally endoscopic approach via RAMT, 86 fulfilled all prespecified inclusion criteria and were subsequently randomized in a 1:1 ratio between the two treatment groups, resulting in 43 patients in the conventional suturing (CS) group and 43 patients in the RAM device (RAM) group.

### Surgical Techniques

All patients were positioned supine with defibrillator pads applied; general anesthesia used single-lumen endotracheal intubation with continuous transesophageal echocardiography (TEE) monitoring. Peripheral cannulation was established percutaneously (Seldinger technique, ultrasound-guided): a dual-stage venous cannula (23–25 Fr) was advanced via the right femoral vein into the superior vena cava, and an 18-Fr arterial cannula was placed in the right femoral artery; in patients weighing >80 kg, additional cannulation of the right internal jugular vein was performed. Correct positioning was confirmed by TEE before CPB.

Thoracic access was obtained through a 3–4 cm incision in the second or third right intercostal space, serving exclusively as the instrument and implant port; a soft tissue retractor (ValveGate™, Geister, Germany) replaced the rib spreader, avoiding rib resection or retraction while preserving the right internal thoracic artery. Two additional 5-mm ports housed a 3D camera (Aesculap EinsteinVision, Tuttlingen, Germany) and a transthoracic Chitwood cross-clamp (Scanlan International, Saint Paul, MN, USA). All steps were performed entirely endoscopically under 3D visualization, without direct vision at any point, the defining feature of our totally endoscopic program, with the surgical image displayed on a 3D monitor opposite the operating surgeon.

After longitudinal pericardiotomy with phrenic nerve preservation, left ventricular decompression used a vent catheter through the right superior pulmonary vein. After mobilization of the proximal AA and aortic cross-clamping, myocardial arrest was induced with Custodiol crystalloid cardioplegia (Dr. Franz Köhler Chemie, Bensheim, Germany), delivered antegrade via the AR or selectively into both coronary ostia in case of significant aortic valve regurtitation.

The dilated AA was resected, preserving a 2-cm distal aortic cuff. In cases needed AR replacement using the Bentall technique, the aortic sinuses were also resected, coronary ostia mobilized as buttons, and native cusps excised.

Randomization determined the technique of annular suture placement and its transfer to the prosthetic sewing cuff, while knot fixation was performed identically in both study arms. In the CS group, annular sutures were manually placed as pledget-reinforced 2-0 polyester mattress sutures and manually passed through the prosthetic sewing cuff. In the RAM group, annular suturing used the automated RAM® COR-SUTURE® QUICK LOAD® device (LSI Solutions, Victor, NY, USA), deployed sequentially right- to non- to left-coronary cusp, delivering double-bite mattress sutures via dual curved needles (3.5/5-mm, sized to the annulus; Figure 1A) and threaded through the sewing cuff using the Sew-Easy system (Figure 1B). In both groups, suture fixation was completed using the Cor-Knot Mini device (Figure 1C), such that RAM and Sew-Easy remained the sole experimental variable between groups. The stepwise use of the RAM device is shown in Supplementary Video S1.

**Figure 1.**
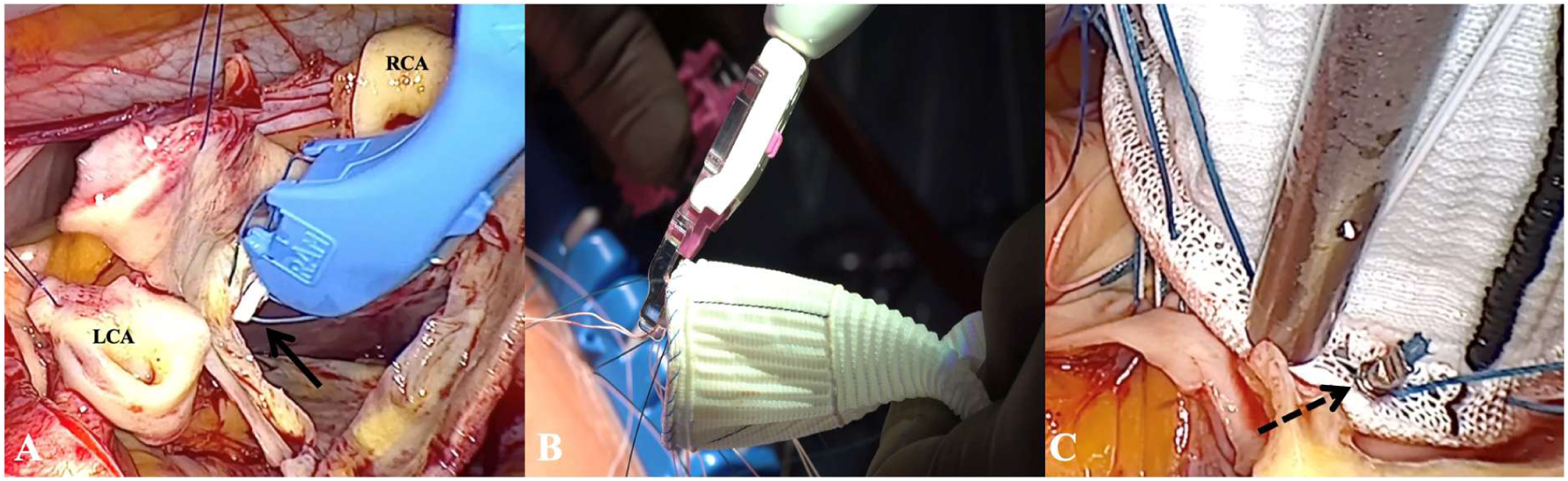
Automated Annular Suturing Technique Using the RAM Device. (A) Placement of the annular suture using the RAM device (LSI Solutions) beginning at the right coronary cusp; the black arrow indicates the pledgeted suture. (B) The Sew-Easy cassettes are loaded onto the Sew-Easy device, guiding suture passage through the sewing cuff of the conduit to complete the horizontal mattress sutures. (C) The Cor-Knot Mini device secures the annular sutures by crimping a titanium fastener onto each pledget while automatically trimming the excess suture material; the black dashed arrow indicates the titanium fastener. CS: conventional suturing group; LCC: left coronary cusp; RAM: automated suturing using the RAM device; RCC: right coronary cusp.

In concomitant AAR and AVR, a bioprosthetic valve was implanted in standard subannular position. In Bentall cases, a valved conduit was fashioned from a bioprosthesis and a Vascutek Valsalva vascular graft (Vascutek Terumo Inc., Inchinnan, Renfrewshire, Scotland, UK) using continuous 3-0 polypropylene suture, or, in anatomically selected patients, a prefabricated biological conduit (KONECT RESILIA™; Edwards Lifesciences, Irvine, CA, USA) was used at the surgeon’s discretion.

For isolated AAR with concomitant AVR, the distal anastomosis (4-0 or 3-0 polypropylene) was performed first, followed by the proximal anastomosis after graft-length determination. In Bentall cases, following conduit parachuting and annular fixation, the left coronary button was reimplanted first (5-0 polypropylene), the distal anastomosis was then completed after graft-length determination, and the right coronary button was reimplanted last (5-0 polypropylene).

After epicardial lead placement and de-airing under TEE guidance, patients were weaned from CPB, with valve function and ventricular/coronary perfusion confirmed by TEE.

After decannulation and femoral closure, pericardial and pleural drains were placed, and the chest was closed in layers with running subcuticular skin closure.

### Postoperative Care

All patients followed a standardized fast-track protocol, applied identically in both groups: early extubation (targeted in the operating room where feasible), multimodal analgesia (intravenous metamizole and piritramide on demand, subcutaneous bupivacaine infiltration), and early mobilization to the bedside within hours and to standing by postoperative day 1.^10^

Patients with intraoperative aortic tissue fragility received light sedation for 1–2 hours before extubation for hemodynamic stabilization.

Patients were admitted to the intensive care unit (ICU) and transferred to intermediate care once extubated and hemodynamically stable.

Chest tube output was recorded at 1, 2, 3, 6, and 24 hours; bleeding was considered significant if output exceeded 400 mL/h within 3 hours of ICU admission or re-thoracotomy, and drains were removed on postoperative day 2 if output was <300 mL/24 h.

Collected variables included cross-clamp and CPB times, ventilation duration, ICU and hospital length of stay, in-hospital and 30-day mortality, reoperations, and major adverce cardiac events (MACE), and secondary outcomes of delirium, pneumonia, re-intubation, acute kidney injury, and permanent pacemaker implantation.

### Endpoints

The primary endpoints of the study were technical success of the procedure, total skin-to-skin, CPB, and aortic cross-clamp times.

Secondary endpoints included in-hospital mortality, 30-day mortality, stroke, conversion to sternotomy, re-thoracotomy for bleeding, paravalvular leakage, duration of mechanical ventilation, length of ICU stay and time to hospital discharge.

### Statistical Analysis

Continuous variables are presented as mean ± standard deviation (SD) or median with interquartile range (IQR), depending on variable distribution. Categorical variables are expressed as absolute numbers and percentages. Continuous variables were tested for normality using the Shapiro–Wilk test and compared using Student’s t-test or Mann–Whitney U test, as appropriate. Categorical variables were analyzed using Pearson χ² test or Fisher exact test. Statistical significance was defined as P < 0.05. All statistical analyses were conducted using IBM SPSS Statistics version 29 (IBM Corp., Armonk, NY, USA) and GraphPad Prism version 8.4.3 (GraphPad Software, La Jolla, CA, USA).

## RESULTS

### Study Population

Baseline demographic, echocardiographic, and comorbidity data are presented in Table 1. Median age was 59.0 years (IQR: 53.0–64.5) in the RAM group and 62.0 years (IQR: 55.5–67.5) in the CS group (P = 0.148), with no significant difference in body mass index or EuroSCORE II. Cardiovascular comorbidities (hypertension, hyperlipidemia, diabetes, coronary artery disease) were comparable between both groups.

**Table 1.** Baseline Data.

|  | Total<br>(n=86) | RAM<br>(n=43) | CS<br>(n=43) | p-value |
| --- | --- | --- | --- | --- |
| Age (year); median; IQR | 60.0 [54.0-65.0] | 59.0 [53.0-64.5] | 62.0 [55.5-67.5] | 0.148 |
| Male sex, n (%) | 67 (79.9%) | 36 (83.7%) | 31 (72.1%) | 0.194 |
| BMI (kg/m<sup>2</sup>); mean ± SD | 26.9±4.21 | 26.8±4.0 | 27.0±4.5 | 0.825 |
| EuroSCORE II (%); median [IQR] | 2.2 [1.8-2.7] | 2.2 [1.8-2.5] | 2.2 [1.9-2.8] | 0.255 |
| **Valve pathology** |  |  |  |  |
| Stenosis, n (%) | 39 (45.3%) | 17 (39.5%) | 22 (51.2%) | 0.379 |
| Regurgitation, n (%) | 9 (10.5%) | 5 (11.6%) | 4 (9.3%) | 0.725 |
| Combined pathology, n (%) | 38 (44.2%) | 21 (48.8%) | 17 (39.5%) | 0.385 |
| Bicuspid valve, n (%) | 55 (64%) | 32 (74.4%) | 23 (53.5%) | **0.043** |
| **Left ventricular ejection fraction** |  |  |  |  |
| >50%, n (%) | 84 (97.7%) | 43 (100%) | 41 (95.3%) | 0.152 |
| 30%-50%, n (%) | 2 (2.3%) | 0 (0%) | 2 (4.7%) | 0.494 |
| **Comorbidities** |  |  |  |  |
| Hypertension, n (%) | 28 (32.6%) | 15 (34.9%) | 13 (30.2%) | 0.645 |
| Hyperlipidemia, n (%) | 28 (32.6%) | 12 (27.9%) | 16 (37.5%) | 0.357 |
| Diabetes, n (%) | 5 (5.8%) | 4 (9.3%) | 1 (2.3%) | 0.360 |
| Smoker, n (%) | 15 (17.4%) | 6 (14%) | 9 (20.9%) | 0.394 |
| Coronary artery disease, n (%) | 10 (11.6%) | 4 (9.3%) | 6 (14%) | 0.738 |
| Chronic obstructive pulmonary, disease, n (%) | 1 (1.2%) | 1 (2.3%) | 0 (0%) | >0.999 |
| Atrial fibrillation, n (%) | 10 (11.6%) | 7 (16.3%) | 3 (7%) | 0.313 |
| Peripheral vascular disease, n (%) | 2 (2.3%) | 1 (2.3%) | 1 (2.3%) | >0.999 |
| Cerebrovascular disease, n (%) | 4 (4.7%) | 1 (2.3%) | 3 (7%) | 0.616 |
| Previous stroke, n (%) | 1 (1.2%) | 0 (0%) | 1 (2.3%) | >0.999 |
| Chronic renal failure, n (%) | 1 (1.2%) | 0 (0%) | 1 (2.3%) | >0.999 |
| **NYHA class** |  |  |  |  |
| I, n (%) | 4 (4.7%) | 2 (4.7%) | 2 (4.7%) | >0.999 |
| II, n (%) | 56 (65.1%) | 28 (65.1%) | 28 (65.1%) | >0.999 |
| III, n (%) | 26 (30.2%) | 13 (30.2%) | 13 (30.2%) | >0.999 |

BMI: body mass index; EuroScore II: European System for Cardiac Operative Risk
Evaluation II; IQR: interquartile range; NYHA: New York Heart Association; PVD:
peripheral vascular disease; SD: standard deviation.

Valve pathology was similar between groups: isolated stenosis in 39 (45.3%), isolated regurgitation in 9 (10.5%), and combined stenosis-regurgitation in 38 (44.2%) patients. In patients with aortic regurgitation, valve-preserving reconstruction was not always feasible due to unfavorable leaflet morphology: cusp thickening/retraction (n = 2), fenestrations (n = 4), cusp sclerosis (n = 3). A bicuspid aortic valve morphology was more frequent in the RAM group (74.4% vs. 53.5%; P = 0.043). Left ventricular ejection fraction exceeded 50% in nearly all patients, and NYHA class II predominated in both groups (65.1%).

### Intraoperative Outcomes

Intraoperative timing outcomes are illustrated in Figure 2. Automated annular suturing showed shorter median total procedure time (120.8 [IQR: 112.0–148.5] vs. 144.0 [IQR: 127.0–175.5] minutes; P = 0.004). CPB duration (90.0 [IQR: 82.5–108.5] vs. 105.0 [IQR: 89.5–121.0] minutes; P = 0.005) and aortic cross-clamp time (62.0 [IQR: 58.0–80.0] vs. 74.0 [IQR: 62.5–89.0] minutes; P = 0.003) were similarly lower in the RAM group. No patient in either group required circulatory arrest for the distal anastomosis.

**Figure 2.**
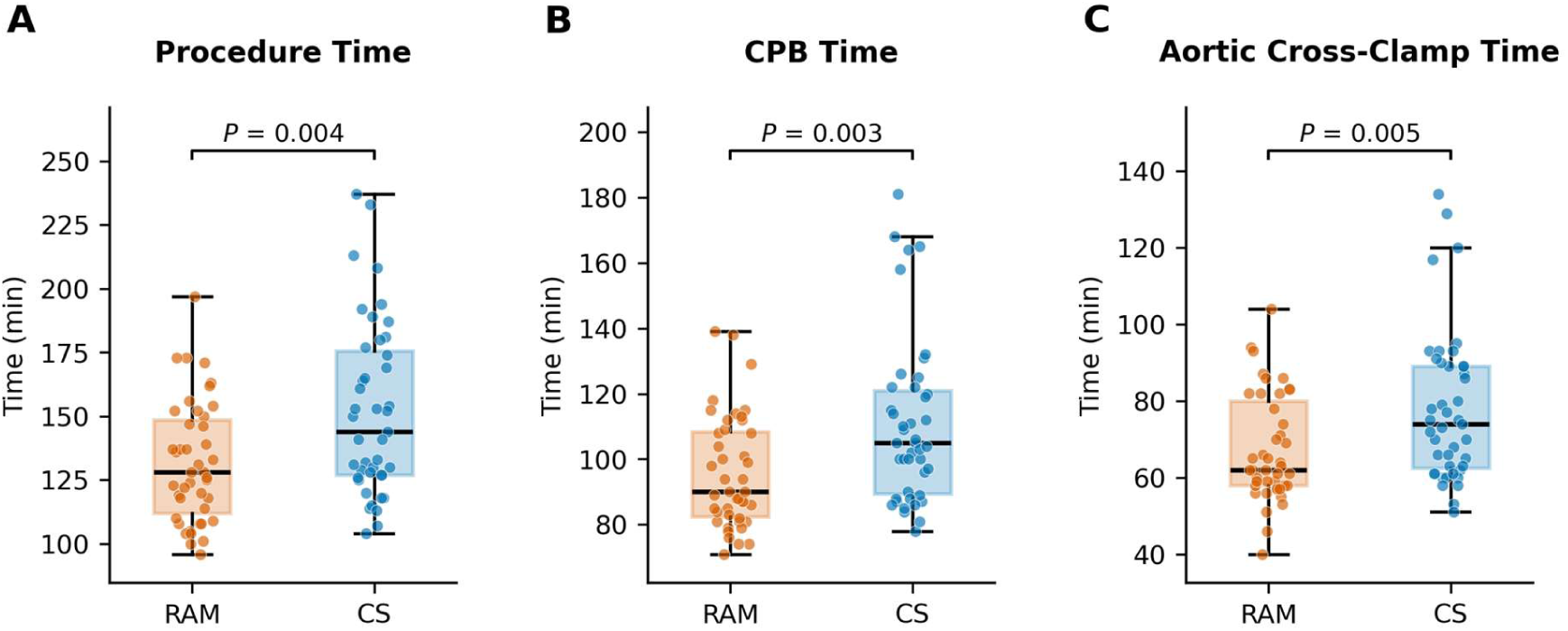
Comparison of intraoperative time metrics between the RAM and CS groups. **(A)** Procedure time (skin-to-skin). **(B)** Cardiopulmonary bypass (CPB) time. **(C)** Aortic cross-clamp time. Box plots show the median (horizontal line) and interquartile range (box); whiskers extend to the most extreme value within 1.5× the interquartile range. Individual dots represent per-patient values (n = 43 per group). CS: Conventional suturing group; RAM: Automated suturing using the RAM device.

### Postoperative Outcomes

Postoperative outcomes are presented in Table 2. Adverse events were uncommon and similar between both arms. Re-thoracotomy for bleeding occurred in one patient per group (2.3%; P > 0.05), while conversion to sternotomy, disabling stroke, acute kidney injury, myocardial infarction, wound infection, and in-hospital or 30-day mortality were zero in both groups. Rates of delirium, pneumonia, re-intubation, and permanent pacemaker implantation were low and did not differ significantly (Table 2).

**Table 2.**
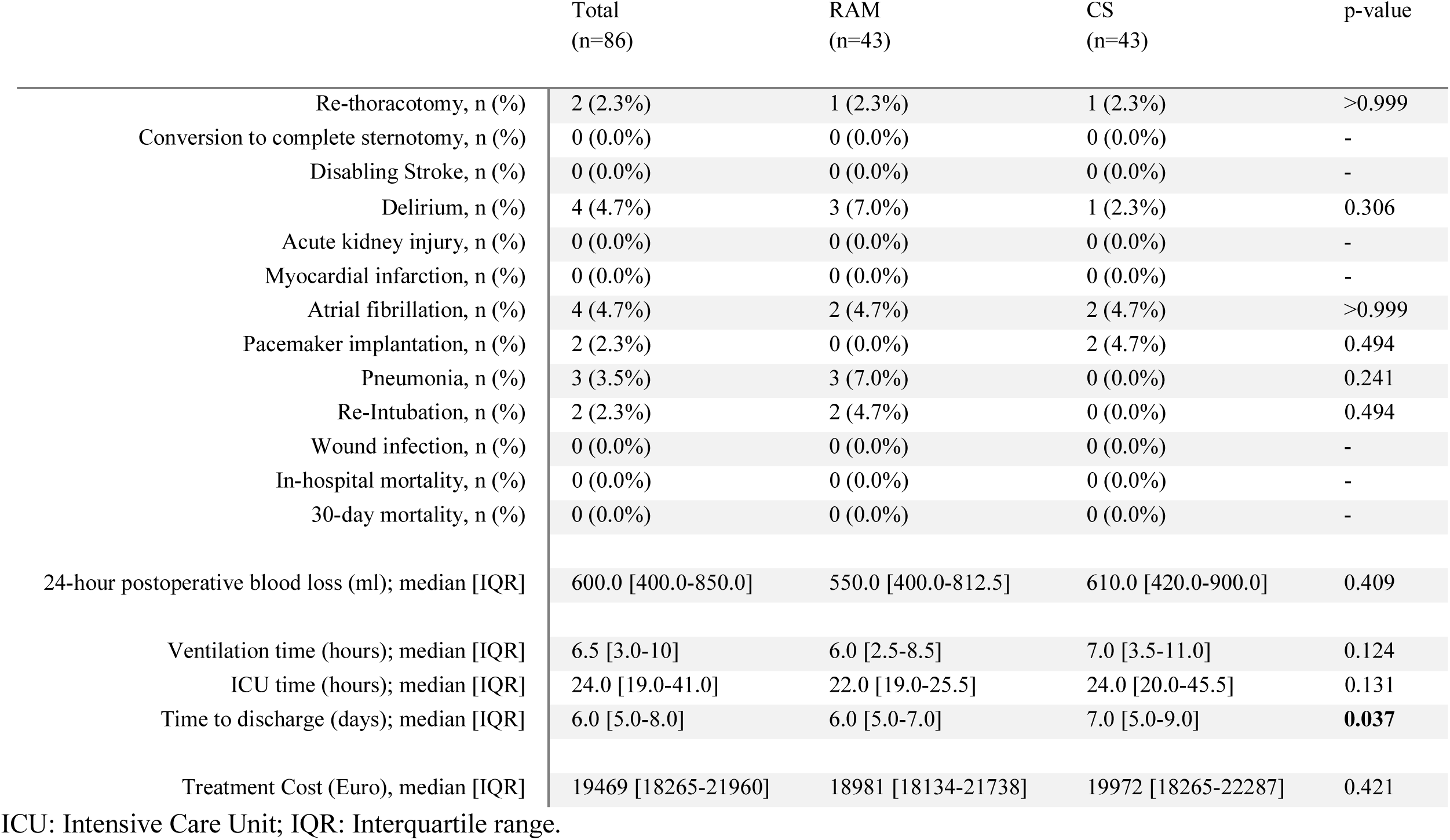
Postoperative Outcome.

|  | Total<br>(n=86) | RAM<br>(n=43) | CS<br>(n=43) | p-value |
| --- | --- | --- | --- | --- |
| Re-thoracotomy, n (%) | 2 (2.3%) | 1 (2.3%) | 1 (2.3%) | >0.999 |
| Conversion to complete sternotomy, n (%) | 0 (0.0%) | 0 (0.0%) | 0 (0.0%) | - |
| Disabling Stroke, n (%) | 0 (0.0%) | 0 (0.0%) | 0 (0.0%) | - |
| Delirium, n (%) | 4 (4.7%) | 3 (7.0%) | 1 (2.3%) | 0.306 |
| Acute kidney injury, n (%) | 0 (0.0%) | 0 (0.0%) | 0 (0.0%) | - |
| Myocardial infarction, n (%) | 0 (0.0%) | 0 (0.0%) | 0 (0.0%) | - |
| Atrial fibrillation, n (%) | 4 (4.7%) | 2 (4.7%) | 2 (4.7%) | >0.999 |
| Pacemaker implantation, n (%) | 2 (2.3%) | 0 (0.0%) | 2 (4.7%) | 0.494 |
| Pneumonia, n (%) | 3 (3.5%) | 3 (7.0%) | 0 (0.0%) | 0.241 |
| Re-Intubation, n (%) | 2 (2.3%) | 2 (4.7%) | 0 (0.0%) | 0.494 |
| Wound infection, n (%) | 0 (0.0%) | 0 (0.0%) | 0 (0.0%) | - |
| In-hospital mortality, n (%) | 0 (0.0%) | 0 (0.0%) | 0 (0.0%) | - |
| 30-day mortality, n (%) | 0 (0.0%) | 0 (0.0%) | 0 (0.0%) | - |
| 24-hour postoperative blood loss (ml); median [IQR] | 600.0 [400.0-850.0] | 550.0 [400.0-812.5] | 610.0 [420.0-900.0] | 0.409 |
| Ventilation time (hours); median [IQR] | 6.5 [3.0-10] | 6.0 [2.5-8.5] | 7.0 [3.5-11.0] | 0.124 |
| ICU time (hours); median [IQR] | 24.0 [19.0-41.0] | 22.0 [19.0-25.5] | 24.0 [20.0-45.5] | 0.131 |
| Time to discharge (days); median [IQR] | 6.0 [5.0-8.0] | 6.0 [5.0-7.0] | 7.0 [5.0-9.0] | **0.037** |
| Treatment Cost (Euro), median [IQR] | 19469 [18265-21960] | 18981 [18134-21738] | 19972 [18265-22287] | 0.421 |

ICU: Intensive Care Unit; IQR: Interquartile range.

24-hour chest tube output was similar between groups (RAM: 550.0 [IQR: 400.0–812.5] vs. CS: 610.0 [IQR: 420.0–900.0] mL; P = 0.409), as were mechanical ventilation duration (RAM: 6.0 [IQR: 2.5–8.5] vs. CS: 7.0 [IQR: 3.5–11.0] hours; P = 0.124) and ICU stay (RAM: 22.0 [IQR: 19.0–25.5] vs. CS: 24.0 [IQR: 20.0–45.5] hours; P = 0.131). Hospital stay was shorter with automated suturing (6.0 [IQR: 5.0–7.0] vs. 7.0 [IQR: 5.0–9.0] days; P = 0.037).

No paravalvular leakage was detected on predischarge echocardiography in either group.

Total treatment costs (Figure 3) were comparable (RAM: €18,981.85 [IQR: 18,134.79–21,738.62] vs. CS: €19,972.84 [IQR: 18,265.20–22,287.66]; P = 0.421).

**Figure 3.**
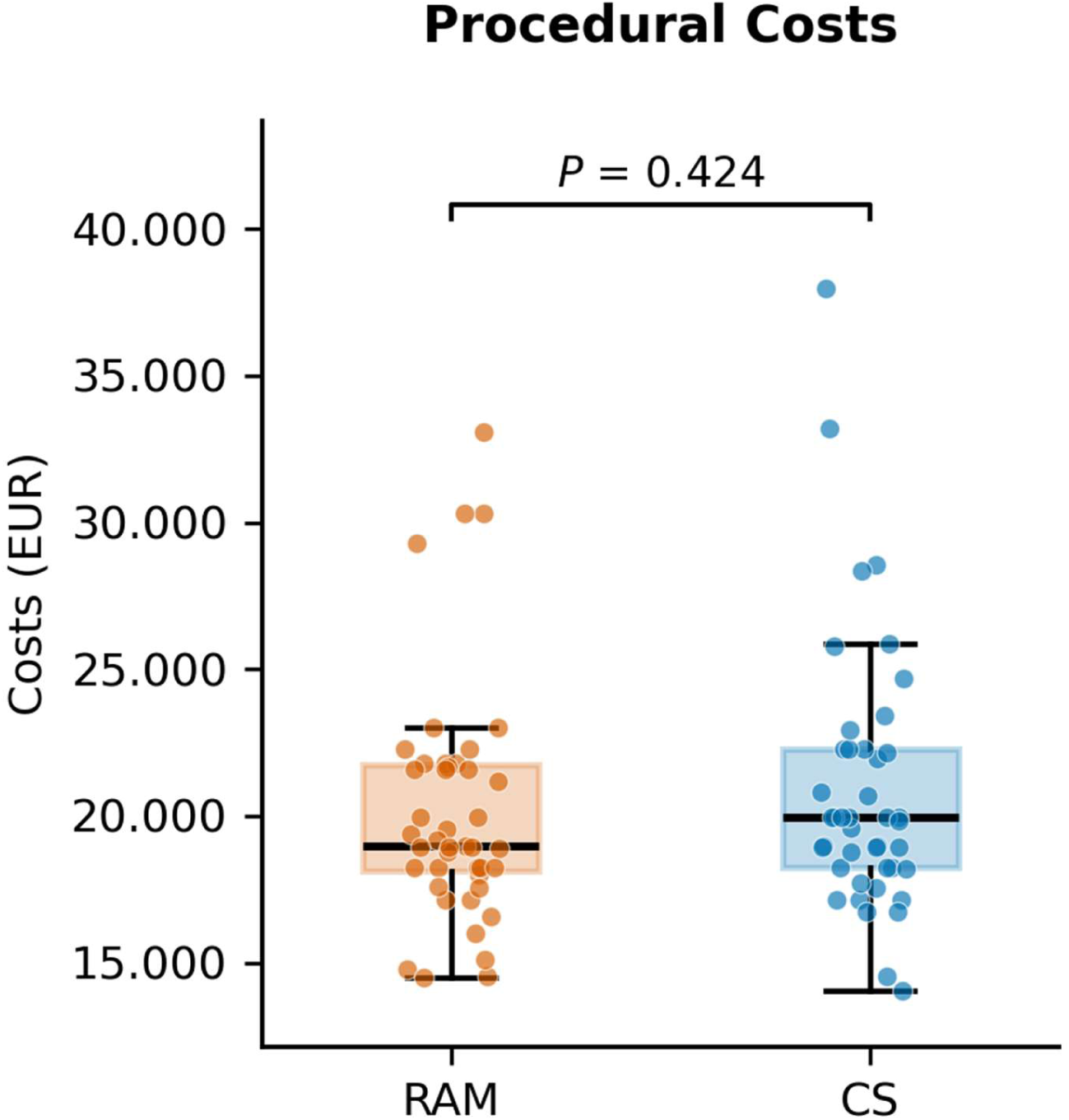
Procedural costs by treatment group. Box plots show the median (horizontal line) and interquartile range (box) of procedural costs (in EUR) for the RAM and CS groups; whiskers extend to the most extreme value within 1.5× the interquartile range. Individual dots represent per-patient costs (n = 43 per group). CS: conventional suturing group; RAM: automated suturing using the RAM device.

## DISCUSSION

This prospective randomized trial demonstrates that automated annular suturing with the RAM device meaningfully improves procedural efficiency during totally endoscopic AVR combined with AA or AR replacement, without compromising safety or surgical quality.

Automated suturing was associated with median reductions of approximately 23 minutes in total procedure time (120.8 vs. 144.0 minutes), 15 minutes in CPB time (90.0 vs. 105.0 minutes), and 12 minutes in aortic cross-clamp time (62.0 vs. 74.0 minutes) compared with conventional suturing. These savings carry particular weight in endoscopic surgery combined with AA or AR replacement, a setting in which limited working space and restricted visualization already narrow the technical margin of the surgeon, and in which CPB and cross-clamp times are inherently longer than in isolated valve surgery owing to the additional aortic steps involved.^5^

Minimally invasive access via RAMT has been linked to less postoperative pain and faster recovery than sternotomy.^11,12^ These benefits are easily eroded when manual suturing extends operative and CPB time; prolonged myocardial ischemic and CPB duration have repeatedly been associated with increased perioperative morbidity, including renal dysfunction, respiratory complications, bleeding, and mortality.^2,13^ Our findings suggest that automated annular suturing may ease one of the most technically demanding components of totally endoscopic valve surgery: by standardizing the spacing and depth of annular stitch placement, the RAM device may improve procedural reproducibility and narrow technical variability between different surgeons. The time saved through automated suturing is therefore concentrated precisely where cumulative procedural risk is highest, suggesting that the RAM device functions less as a marginal efficiency gain than as a targeted mitigation of the specific time burden introduced by combining AVR and AAR within a single totally endoscopic procedure.

Mechanistically, the RAM system places both needles of each suture simultaneously at a fixed, predefined distance (3.5 or 5 mm) within the annular tissue; sutures were then transferred through the prosthetic sewing cuff at identical spacing using the Sew-Easy device in the RAM group, whereas this step was performed manually in the conventional group. Suture fixation in both arms was completed with the Cor-Knot titanium fastener, held constant across groups.^14^ The differences in procedural efficiency observed in this trial can accordingly be linked to the combined effect of automated annular suture placement and automated prosthetic suture transfer (RAM and Sew-Easy) rather than to the knot-fastening step itself.

The efficiency gained with automated suturing did not appear to come at the expense of safety: no differences in stroke, paravalvular leakage, or conversion to sternotomy were observed between groups. Paravalvular leakage typically arises from incomplete apposition of the sewing ring to native tissue due to suture dehiscence or restricted access to the annular plane; the fixed inter-suture spacing enforced by the RAM and Cor-Knot systems may help mitigate this mechanism, a possibility consistent with its absence in either arm of our cohort.^15^

The significantly shorter hospital stay observed with automated suturing likely reflects the cumulative effect of reduced operative trauma and shorter myocardial ischemic time; ICU stay and ventilation duration, while not significantly different, trended numerically in the same direction.

Despite this reduction in length of hospital stay, total treatment cost did not differ significantly between both study arms (RAM: €18,981.85 vs. CS: €19,972.84; P = 0.421), a finding that merits direct comment, as it runs counter to the intuitive expectation that a shorter admission should lower overall cost. Disaggregating material costs clarifies this result: the Cor-Knot fastening system was applied identically in both study arms at a fixed cost of €658.33 per case and therefore cancels out in the between-group comparison, while the RAM device and Sew-Easy cassette, used exclusively in the automated group, added an incremental €1,798.96 per case. This device-specific increment largely offset the savings otherwise attributable to shorter length of hospital stay, rendering automated suturing effectively cost-neutral rather than cost-saving from the economic perspective in the present analysis. We view this as a point of transparency rather than a limitation: institutions considering adoption of automated annular suturing should not expect the acquisition cost to be recovered through reduced length of hospital stay alone within the index hospitalization. Our analysis was restricted to direct per-case treatment cost and does not capture potential indirect institutional benefits, such as shorter operating room occupancy enabling higher case throughput, or downstream cost avoidance from a reduced burden of paravalvular leakage–related reintervention. Whether these effects would shift the overall economic balance over a longer time horizon remains an open question for future health-economic analysis.

These findings are consistent with prior experience using automated annular suturing in isolated totally endoscopic AVR, extending the observation to the technically more demanding setting of combined AVR with AA or AR replacement.^16–18^

Combined AVR and AA or AR replacement via a totally endoscopic approach remains among the most technically demanding procedures in minimally invasive cardiac surgery, given restricted visualization, limited instrument maneuverability, and the need for efficient myocardial protection during prolonged reconstructive phases. By enabling rapid, reproducible annular suture placement, the RAM device may help address one of the principal technical barriers to broader adoption of totally endoscopic aortic surgery, potentially shortening the learning curve and reducing interoperator variability.^12,19,20^ No increase in procedure-related complications accompanied its use, including a low, statistically similar rate of permanent pacemaker implantation, supporting the safety of this approach without evidence of increased conduction disturbance.

This study reflects the broader trajectory of totally endoscopic cardiac surgery toward increasingly complex procedures. As endoscopic techniques extend from isolated valve interventions to combined aortic surgery, enabling technologies such as automated suturing are likely to play a growing role in preserving procedural reproducibility and efficiency as case complexity rises.

### LIMITATIONS

Several limitations warrant consideration. This single-center randomized trial had a moderate sample size and was performed at a high-volume center experienced in totally endoscopic aortic surgery; confirmation in larger, multicenter cohorts, including centers earlier in their learning curve, is needed to establish generalizability. Follow-up was limited to the early postoperative period; longer-term data are needed to assess prosthetic durability, late paravalvular leakage, endocarditis, and survival. Annular suturing time was not isolated as a discrete variable, and a higher proportion of bicuspid valves in the RAM group, together with increasing surgeon experience, cannot be fully disentangled from the device effect; nonetheless, all procedures were performed by experienced minimally invasive surgeons, with automated suture placement and transfer the principal difference between groups. Finally, our cost analysis was restricted to per-case direct costs; institutional-level effects, such as increased operating room throughput, remain speculative.

## CONCLUSION

In this randomized trial, automated annular suturing with the RAM device significantly reduced total procedure, CPB, and aortic cross-clamp times during totally endoscopic AVR with concomitant AAR, without any signal of compromised safety. This efficiency translated into a shorter hospital stay, though not into lower overall treatment cost once the incremental device expense was accounted for. Automated annular suturing represents a safe and effective enabler of increasingly complex totally endoscopic aortic procedures.

## Data Availability

The data, analytic methods, and study materials will be made available to other researchers upon reasonable request to the corresponding author for purposes of reproducing the results or replicating the procedure.

## Acknowledgment

The authors sincerely thank the patients who voluntarily participated in this study for their trust and cooperation.

## Sources of Funding

The authors have no internal or external funding sources to disclose.

## Disclosures

Farhad Bakhtiary reports a relationship with Edwards Lifesciences, Medtronic, Corcym, and Abbott that includes: consulting or advisory and speaking fees. Farhad Bakhtiary reports a relationship with LSI that includes: speaking fees. Other authors declare that they have no known competing financial interests or personal relationships that could have appeared to influence the work reported in this paper.

## Non-standard Abbreviations and Acronyms

AA: ascending aorta
AAR: ascending aorta replacement
AR: aortic root
AVR: aortic valve replacement
BMI: body mass index
CPB: cardiopulmonary bypass
CS: conventional suturing
CT: computed tomography
EuroSCORE II: European System for Cardiac Operative Risk Evaluation II
ICU: intensive care unit
IQR: interquartile rang
LCC: left coronary cusp
MACE: major adverse cardiac events
NYHA: New York Heart Association
PVD: peripheral vascular disease
RAM: automated suturing using the RAM device
RAMT: right anterior minithoracotomy
RCC: right coronary cusp
SD: standard deviation
TEE: transesophageal echocardiography

## Notes

### Competing Interest Statement

The authors have declared no competing interest.

### Clinical Trial

German Registry for Clinical Studies, Trial number: DRKS00030520, registry date: 18th December 2023

### Author Declarations

Approval by the institutional ethics committee of the University of Bonn (approval 13 no. 184/23; August 7, 2023)

## REFERENCES

1. Bakhtiary F, Salamate S, Eghbalzadeh K, El-Sayed Ahmad A. Endoscopic micro-invasive cardiac surgery: state-of-the-art. Turk Gogus Kalp Damar Cerrahisi Derg. 2024;32:355–366.

2. Holubec T, Dahle G, Bonaros N. Editorial: minimally invasive cardiac surgery: state of the art and current challenges. Front Cardiovasc Med. 2023;10:1286868.

3. El-Sayed Ahmad A, Salamate S, Bakhtiary F. Lessons learned from 10 years of experience with minimally invasive cardiac surgery. Front Cardiovasc Med. 2022;9:1053572.

4. El-Sayed Ahmad A, Bayram A, Salamate S, Sirat S, Amer M, Bakhtiary F. Percutaneous versus surgical femoral access in minimally invasive cardiac operations. Eur J Cardiothorac Surg. 2022;61:1348–1354.

5. Hamiko M, Salamate S, Nassari MA, et al. Totally endoscopic replacement of the ascending aorta and the aortic root including the aortic valve via right minithoracotomy: a multicenter study. J Clin Med. 2024;13:2648.

6. Jenkin I, Prachee I, Sokal PA, Harky A. The role of Cor-Knot in the future of cardiac surgery: a systematic review. J Card Surg. 2020;35:2987–2994.

7. Bakhtiary F, Salamate S, Ahmad AE. Totally endoscopic micro-invasive aortic valve replacement. Multimed Man Cardiothorac Surg. 2024;2024.

8. Elghannam M, Aljabery Y, Naraghi H, et al. Minimally invasive aortic root surgery: midterm results in a 2-year follow-up. J Card Surg. 2020;35:1484–1491.

9. Czerny M, Grabenwöger M, Berger T, et al. EACTS/STS guidelines for diagnosing and treating acute and chronic syndromes of the aortic organ. Ann Thorac Surg. 2024;118:5–115.

10. Kruse J, Krasny I, Hamiko M, et al. Enhanced recovery after minimally invasive valve surgery: ultra-fast extubation versus standard weaning strategy. Sci Rep. 2025;15:35048.

11. Fatehi Hassanabad A, King MA, Karolak W, et al. Right anterior minithoracotomy approach for aortic valve replacement. Innovations (Phila). 2024;19:494–508.

12. Sazzad F, Ler A, Kuzemczak M, et al. Automated fastener vs hand-tied knots in heart valve surgery: a systematic review and meta-analysis. Ann Thorac Surg. 2021;112:970–980.

13. Goebel N, Bonte D, Salehi-Gilani S, et al. Minimally invasive access aortic arch surgery. Innovations (Phila). 2017;12:351–355.

14. Sazzad F, Xian OZ, Ler A, et al. Incidence of valvular regurgitation and leaflet perforation by using automated titanium fasteners (CORKNOT) in heart valve repair or replacement: less usual than reported. J Cardiothorac Surg. 2021;16:163.

15. García E, Sandoval J, Unzue L, et al. Paravalvular leaks: mechanisms, diagnosis and management. EuroIntervention. 2012;8(suppl Q):Q41–Q52.

16. Risteski P, El-Sayed Ahmad A, Monsefi N, et al. Minimally invasive aortic arch surgery: early and late outcomes. Int J Surg. 2017;45:113–117.

17. Harky A, Al-Adhami A, Chan JSK, et al. Minimally invasive versus conventional aortic root replacement: a systematic review and meta-analysis. Heart Lung Circ. 2019;28:1841–1851.

18. Hamiko M, Salamate S, Bayram A, et al. Feasibility and safety of totally endoscopic Bentall procedure via right anterior mini-thoracotomy: early experience. JTCVS Tech. 2026;36:102277.

19. El-Sayed Ahmad A, Salamate S, Granov N, et al. First experiences with automated annular suturing device in totally endoscopic aortic and mitral valve replacement. Interdiscip Cardiovasc Thorac Surg. 2024;38:ivae112.

20. Pitsis A, Tsotsolis N, Boudoulas H, Boudoulas KD. Totally endoscopic aortic valve replacement with concomitant trans-aortic mitral valve repair for mitral regurgitation. J Cardiothorac Surg. 2021;16:318.

